# Genome-wide association studies on malaria in Sub-Saharan Africa: a scoping review

**DOI:** 10.1101/2024.08.11.24311829

**Authors:** Morine Akoth, John Odhiambo, Bernard Omolo

## Abstract

**Background:** Malaria remains one of the leading causes of death in Sub-Saharan Africa (SSA). The scoping review mapped evidence in research on existing studies on malaria genome-wide association studies (GWAS) in SSA.

**Methods:** A scoping review was conducted to investigate the extent of malaria studies in SSA under GWAS. The review followed the methodology for scoping reviews developed by Arksey and O’Malley, including identification of research problems, searching for relevant studies, selecting studies, charting data, collating, summarizing, and reporting the findings. Data from relevant studies were collected and synthesized using Excel and Zotero software. The data collected included information on the author, the years of study, the countries of study, the research areas of interest, and the study designs used.

**Results:** Of an initial pool of over 2000 articles retrieved from four databases, namely Google Scholar, PubMed, Scopus, and Web of Science, 569 were retained. After applying the inclusion-exclusion criteria, 99 articles were found to be relevant. Most of these studies (n=25, 60%) used a case-control study design, while the rest used cross-sectional, cohort, longitudinal, family-based, and retrospective designs. These studies were conducted between 2000 and 2023, with a significant increase observed in 2011. Most studies were carried out in Kenya (n = 25), Gambia (n = 17), Cameroon (n = 15), Ghana (n = 12), and Tanzania (n=11), primarily exploring genetic variants associated with malaria susceptibility, resistance, and severity.

**Conclusion:** Many case-control studies in Kenya and Gambia reported genetic variants in malaria susceptibility, resistance, and severity. Few articles were systematic reviews and scoping reviews. GWAS on malaria is scarce in SSA and even fewer studies are model-based. Consequently, there is a pressing need for more genome-wide research on malaria in SSA.

## Introduction

Genome-wide association studies (GWAS) are a powerful tool to identify genes linked to human diseases, attracting global attention among researchers [1]. Malaria GWAS is vital among the African population, as the disease continues to be a challenge that takes a heavy toll on human health and socioeconomic development [2–6]. The World Health Organization (WHO) report estimated 599 thousand deaths in the year 2020, and 234 million cases in the year 2021 in Africa, accounting for 95% of the global malaria cases [7, 8]. GWAS have become powerful tools for understanding the genetic basis of complex diseases, including susceptibility, severity, and resistance to malaria [9–12]. Through studying the entire genome for associations between genetic variants and disease phenotypes, GWAS has provided information on the genetic architecture of malaria-related traits, paving the way for the development of targeted interventions and personalized therapies [13]. In the context of SSA, where genetic diversity is particularly high [14] and the malaria burden is felt [9], GWAS holds immense promise for advancing our understanding of malaria epidemiology, pathogenesis, and treatment outcomes.

Most GWAS have been conducted in Europe and Asia [15, 16], and Europe has reported bias in large-scale genomic studies. The international consortia for collaboration of genetic studies have left out Africa due to limited data and resources [17]. This has led many African countries not being represented in the research despite the genetic diversity among African populations [14, 18, 19]. Despite all these, the field of genomics has advanced considerably due to a few current initiatives. Some of these initiatives are 54Gene, The African Centre of Excellence for Genomics of Infectious Diseases (ACEGID), Inqaba Biotec (Africa’s Genomics Company), and The Human Heredity and Health in Africa (H3Africa) Consortium [20]. Ziyaad and colleagues [21], for instance established the *H*3*Africa Archive* for African human genomic data management. This effort aimed to facilitate data sharing and access, addressing the challenge of accessing high-throughput technologies in genomics.

Therefore, the primary objective of this scoping review was to chart the existing GWAS on malaria in the SSA. The databases were systematically searched to identify eligible studies published in English from January 2000 to December 2023. The year of study, study designs, subject areas, and countries of study were mapped and reported. A search strategy was developed to identify relevant literature using the Arksey and O’Malley [22] framework. The search was tailored to four databases: Google Scholar, PubMed, Scopus, and Web of Science. The search terms used were “Genome-wide association studies”, “Malaria resistance”, “Malaria”, “Genetic association testing”, and “Sub-Saharan Africa ”. All searches included journal articles that were peer-reviewed.

## Methods

### Study design

The study design aimed to conduct a comprehensive search for studies on GWAS in malaria within the SSA region, following the framework established by Arksey and O’Malley [22]. The framework has five steps: identifying the research question, identifying relevant studies, selecting studies, charting data, collating, summarizing, and reporting the data.

#### Identifying research question

The main research question for the scoping review was: What are the existing genome-wide association studies on malaria in SSA conducted between 2000 and 2023? Table 1 shows the population, concept, and context guide (PCC) used to assess the eligibility of the research question and to guide the selection of studies derived from the Joanna Briggs Institute [23].

**Table 1.** PCC framework used to determine the eligibility of the research question and to guide the selection of studies on GWAS in malaria.

|  |  |
| --- | --- |
| *Population* | General population (adults and children) |
| *Concept* | Genome-wide association studies in malaria |
| *Context* | Sub-Saharan Africa |

#### Identifying relevant studies

Identifying relevant articles required a comprehensive search from 2000 to 2023 in four databases: Google Scholar, Scopus, PubMed, and Web of Science. These are life science journals with biomedical literature.

#### Search strategy

A methodology was devised to examine the existing GWAS on malaria in SSA. Furthermore, the research sought to identify and investigate model-based studies that addressed the concept of heterosis. We searched the literature on malaria GWAS using the mentioned databases with the last search conducted on January 21, 2024, and included all articles published up to December 31, 2023. A combination of the following keywords was used: “Genome-wide association studies”, “Malaria resistance”, “Malaria”, “Genetic association testing”, and “Sub-Saharan Africa ”. A total of 569 articles were initially retrieved from the four databases. Following the removal of duplicates and irrelevant articles, a screening process was applied to 99 articles. The Preferred Reporting Items for Systematic Reviews and Meta-Analysis extension for scoping reviews (PRISMA-ScR) guidelines were followed to identify, select, and include relevant articles [24].

#### Inclusion and exclusion criteria

The criteria for inclusion of articles in the scoping review were as follows: The study must be a GWAS conducted in SSA between January 2000 and December 2023, and focused on malaria, the study must be published in English, and the study must have the full text available. The exclusion criteria consisted of the following: GWAS conducted outside the SSA region, studies related to malaria that did not fall under the scope of GWAS, studies conducted in the regions of Algeria, Egypt, Morocco, Tunisia, and Libya, studies investigating non-malarial phenotypes and systematic review, meta-analysis, and other reviews.

#### Selecting studies

The titles of selected articles from various repositories were filtered and then identified, and duplicates were removed using Excel software. The primary data extraction tool employed was Microsoft Excel, while Zotero served as the reference management software for extracting and organizing citation details. Microsoft Excel was also used to manage and chart extracted data. Two independent reviewers evaluated the abstracts of qualified articles based on predetermined inclusion and exclusion criteria and resolved any discrepancies. Articles on reviews and systematic studies were recorded but were excluded from the scoping review. The database searches, keywords used, and the number of selected articles were noted.

#### Charting the data

Information was extracted using the Excel software, including author names, publication dates, article titles, study designs, study country, study areas, genetic modes of inheritance for the model-based studies, genetic variants associated with malaria for some studies, and other significant findings. Two independent reviewers performed data extraction and analysis of the selected studies by manually studying full articles.

Quality assessment for studies followed established screening criteria, and for review, the PRISMA-ScR guidelines were applied for quality evaluation.

#### Collating, summarizing and reporting results

Emerging themes related to GWAS in malaria were summarized. A descriptive analysis of peer-reviewed papers published and addressed the research question. The research covered various aspects, including the year of publication, the number of publications in different countries in SSA, the different study designs used, the categorization of subject areas into distinct categories such as susceptibility to malaria, severe malaria, and resistance, the methodologies used to evaluate genetic association, drug resistance, population diversity, and host-parasite interactions. Tables and graphs were employed where appropriate to represent the findings visually. *R* software was used to summarize and categorize the data. *SRplot* platform, a data visualization and graphing tool ([25]), was also utilized to create the figures in compliance with the required standards. The scoping review results were used to identify knowledge gaps on malaria GWAS in SSA. The data and materials linked to this study are now available through the Open Science Framework (OSF) repository at https://doi.org/10.17605/OSF.IO/DFK5G.

## Results

A total of 569 studies were found in four electronic databases (Google Scholar (n=476), PubMed (n=29), Web of Science (n=8), and Scopus (n=56)). After removing 89 duplicate and 370 ineligible records for various reasons (not within the human population, not malaria GWAS and not published in English), 117 unique records underwent title and abstract screening. Of these, 9 records were irrelevant and excluded (4 non-GWAS, 4 non-malaria phenotypes, 1 not from the human population), leaving 108 reports for full-text retrieval. However, five records were either inaccessible or only had abstracts published, leaving 103 full-text reports for eligibility assessment by at least two reviewers. After full-text screening, four more records were excluded and 99 studies were included in this scoping review (Fig 1).

**Fig 1.**
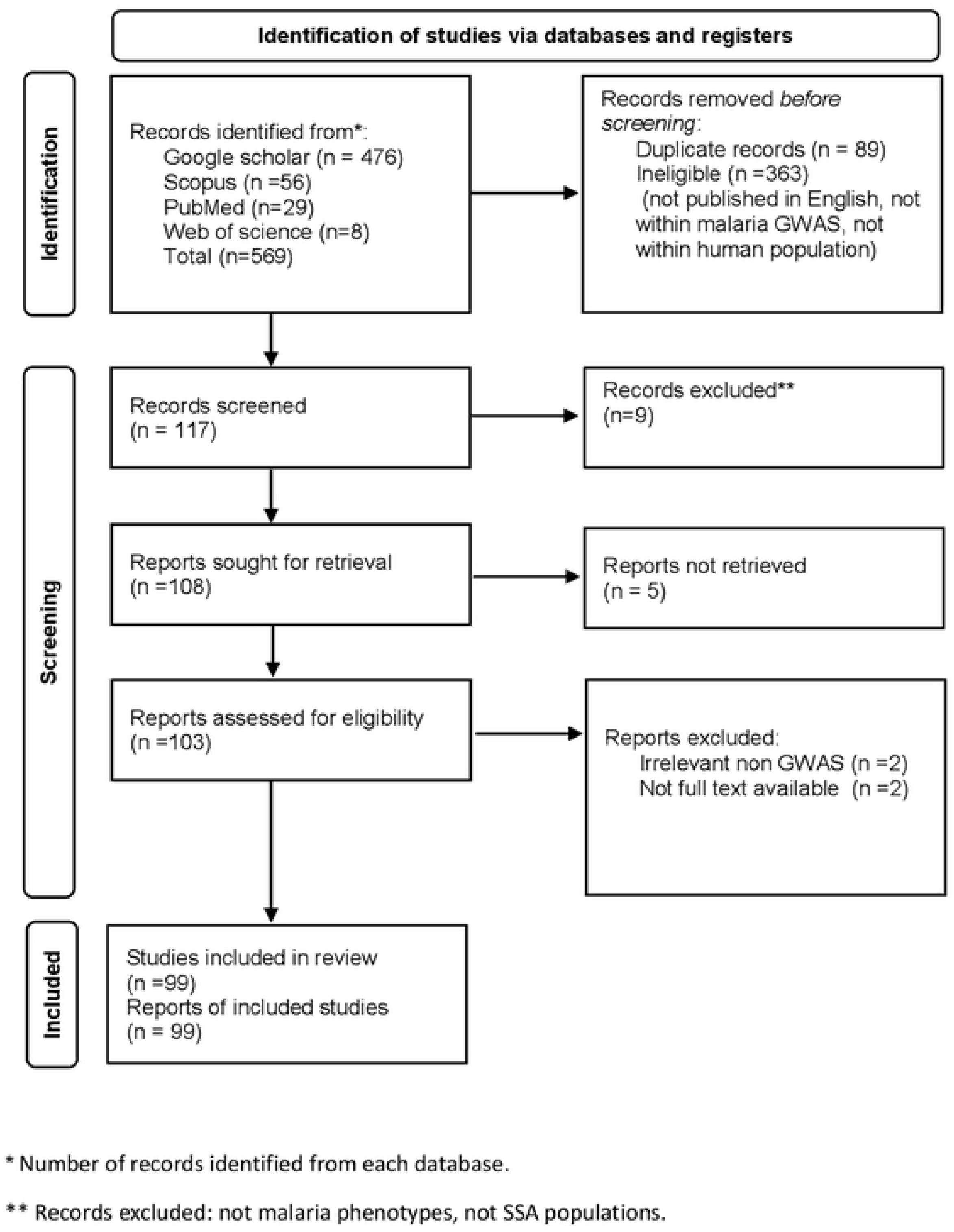
Preferred Reporting Items for Systematic Reviews and Meta-Analysis extension for 64 scoping reviews (PRISMA-ScR) on malaria GWAS in SSA.

**Fig 2.**
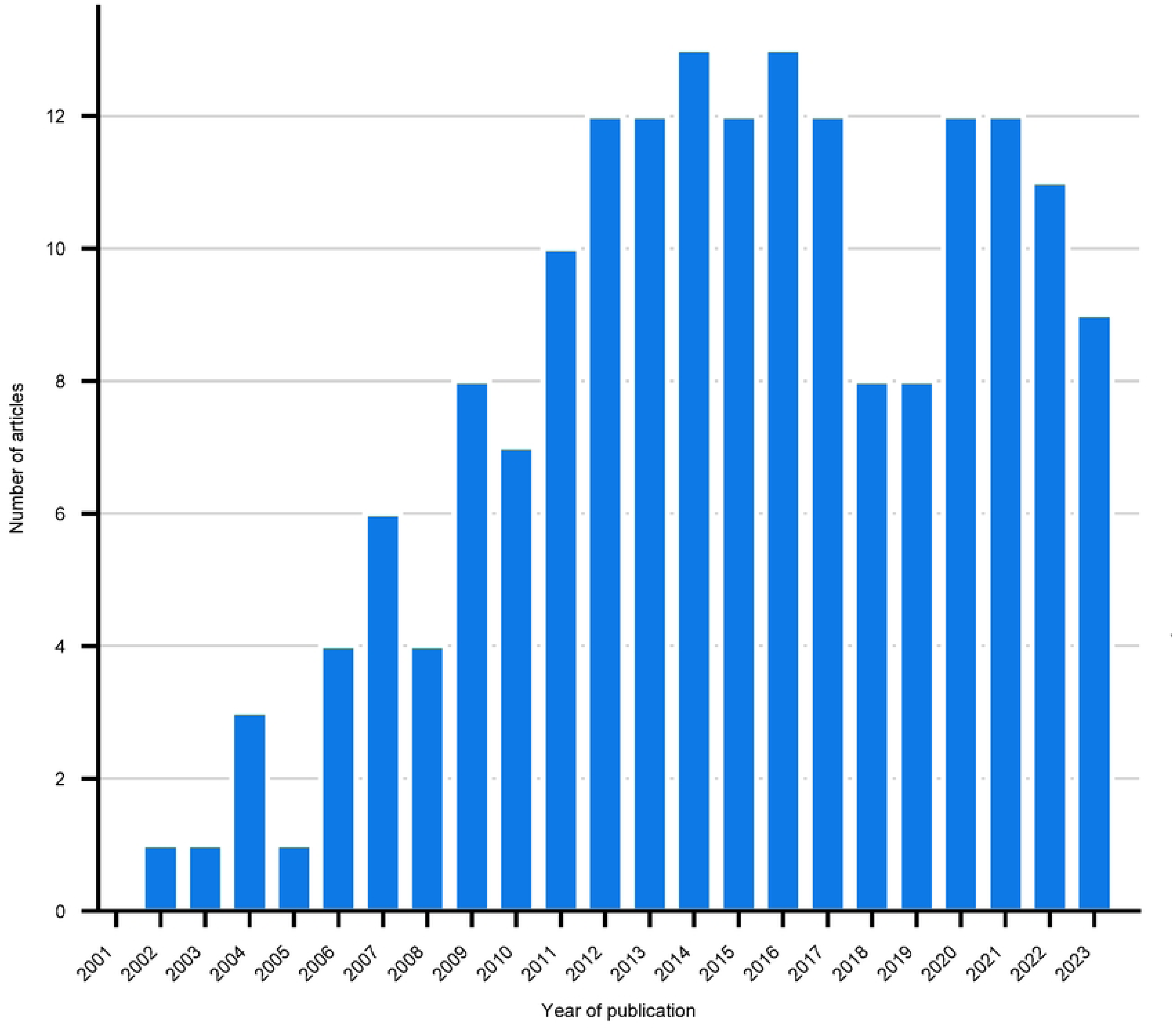
Number of articles in malaria GWAS in SSA published between 2000 and 2023.

### Study designs and areas

The predominant study design among the included investigations was case-control studies (*n* = 25, 60%) [9, 10, 26–50], followed by cohort studies (*n* = 8, 19%) [51–58], then family-based studies (*n* = 6, 14%) [34, 59–61], cross-sectional and longitudinal studies (*n* = 3, 6%) [57, 62, 63]. In the study, systematic reviews (n = 14) [9, 30, 49, 64–69] and reviews (n=7) [16, 70–76], were also reported. Furthermore, there was a retrospective study [77] (Fig 3).

**Fig 3.**
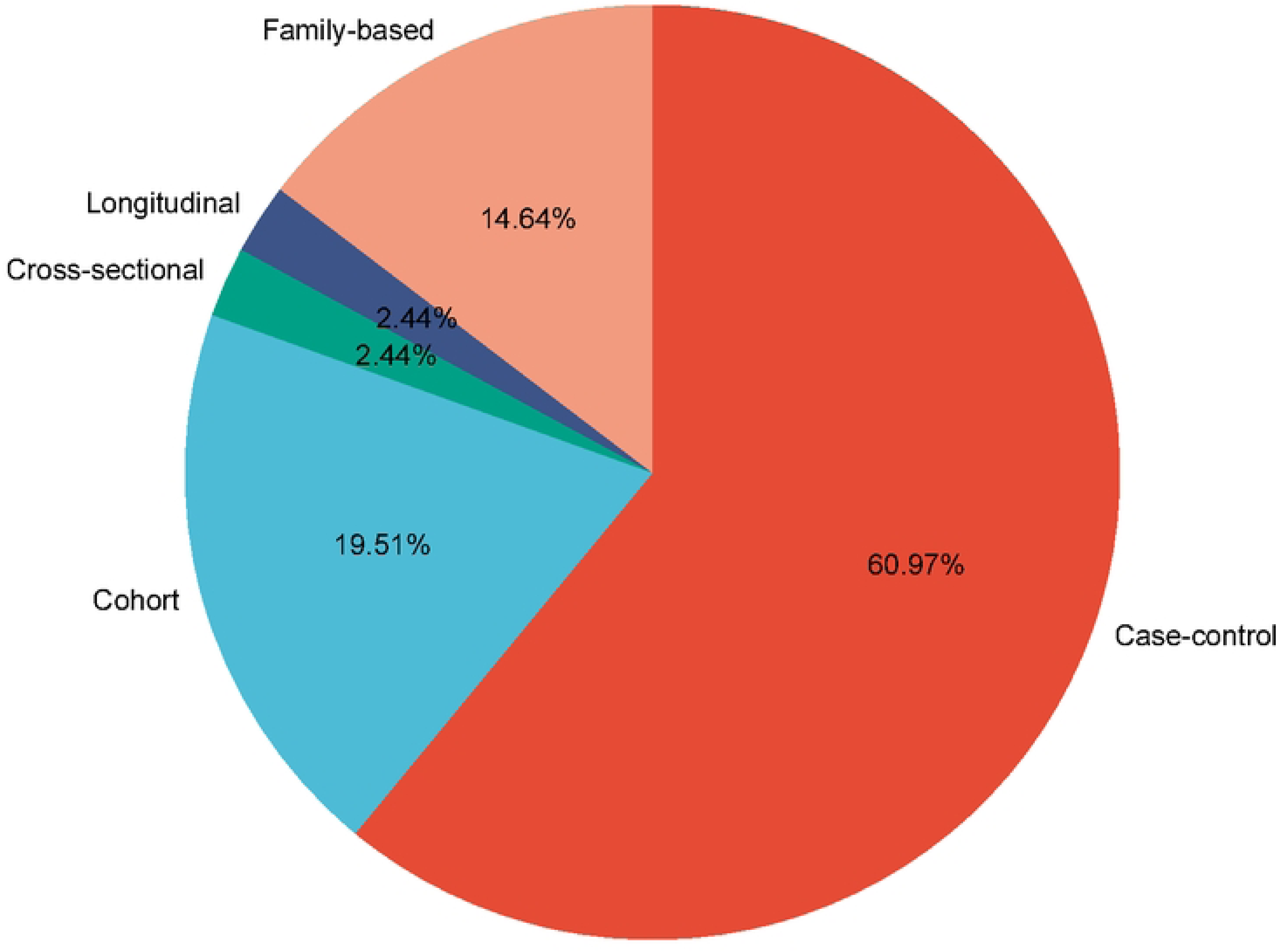
Different study designs used in malaria GWAS.

Most GWAS on malaria focused on identifying genetic variants associated with susceptibility, resistance, and severity of malaria [2, 8–10, 26, 27, 29–38, 41, 42, 44–50, 52, 54–58, 61, 63–65, 67–69, 71, 74, 78–83]. Furthermore, considerable research had been conducted on methodological approaches to genetic association testing, malaria drug resistance patterns exhibited by malaria parasites, and the effectiveness of vaccine interventions [31, 40, 43, 46, 51, 59, 60, 70, 75, 77, 81, 84–100]. Other areas of study included host-parasite interactions, population diversity and structures, genetic variation and evolutionary insights such as gene flow and natural selection [8, 16, 30, 56–58, 61, 68, 71, 101–108] and Mendelian randomization [109] (Fig 4)

**Fig 4.**
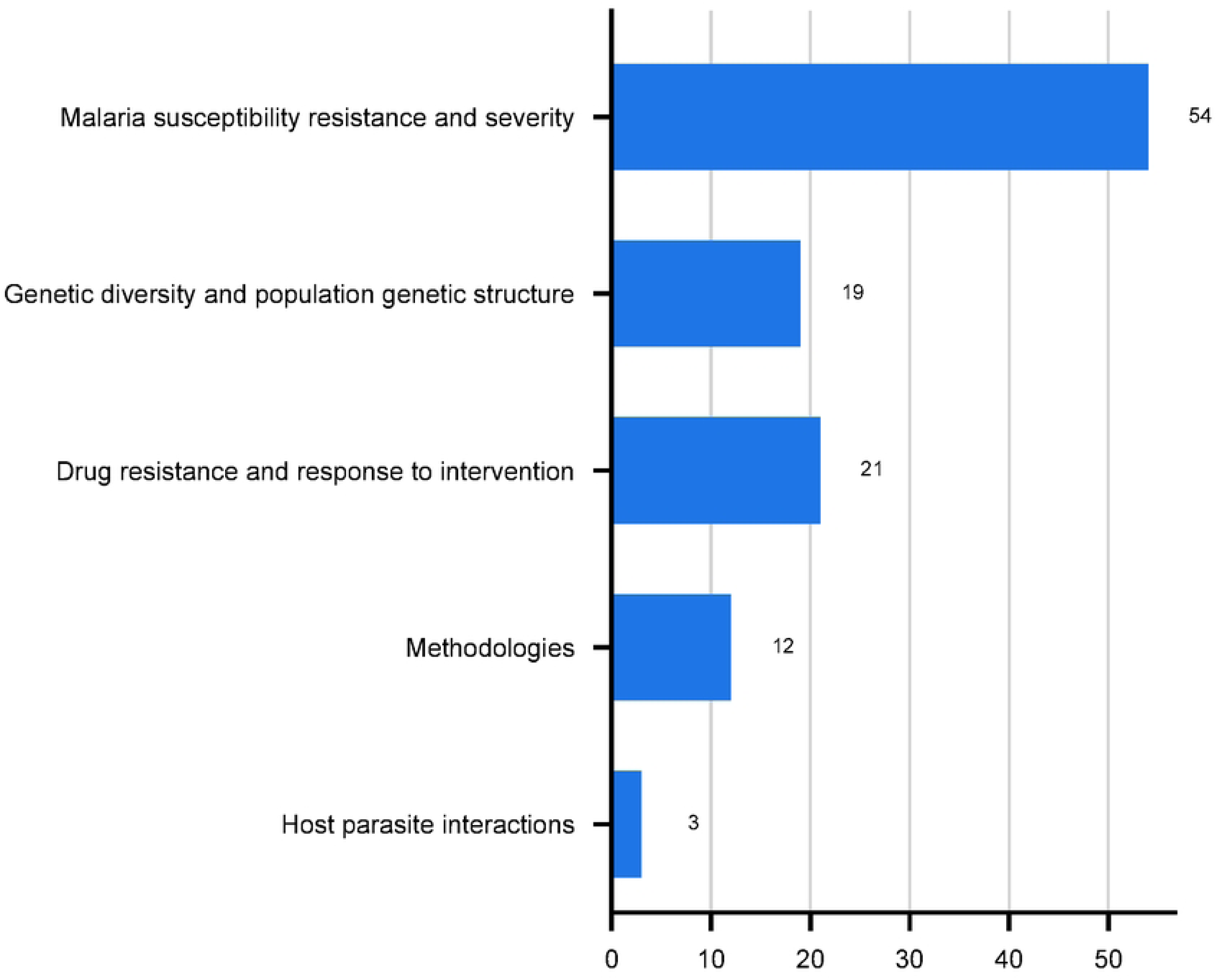
Different research areas in malaria GWAS in SSA.

### Year of study and spatial distribution of studies

Studies reported were published between 2000 and 2023, with a noticeable increase in publications from 2011 as shown in Fig 2.

The study covered 27 different locations across various countries in SSA, with Kenya having the highest number of articles (n=25) [9, 30, 35, 35, 36, 40–42, 46, 49, 50, 56, 64, 71, 80, 83, 85, 87, 91, 93, 98, 109–111] as shown in Fig 5. Other countries included Gambia (n=17) [9, 26, 30, 34, 35, 40, 42, 43, 45, 56, 61, 64, 67, 90], Cameroon (n=15) [9, 28, 35, 51, 53, 71, 109, 112], Tanzania (n=11) [9, 27, 35, 37, 55, 87, 109, 113, 114], Ghana (n=12) [10, 42, 69, 94, 97, 100, 102], Malawi (n=7) [35, 42, 56, 57, 64, 91, 96], Burkina Faso (n=4) [35, 36, 52, 115], Uganda (n=3) [69, 96, 104], South Africa (n=1) [2], Togo (n=3) [94, 100, 105], Mali (n=2) [48, 63], Benin (n=7) [47, 58, 94–96, 99, 100], Ethiopia (n=2) [71, 77], Ivory Coast (n=4) [44, 94, 100, 107], Mozambique (n=1) [39], Nigeria(n=2) [35, 93], Guinea (n=1) [102] Senegal (n=5) [8, 60, 68, 88, 107], Sudan (n=1) [61], Mauritania (n=1) [71], Equatorial Guinea (n=1) [71], Angola (n=1) [71], Democratic republic of Congo (n=1) [107] and Madagascar (n=1) [71]. Some studies covered more than one region of SSA and the research design was mainly systematic reviews and meta-analyses [9, 30, 56, 64, 115]. There were no malaria GWAS in some SSA regions such as Rwanda, Somalia, Chad, Namibia, Sierra Leone, Niger and Burundi.

**Fig 5.**
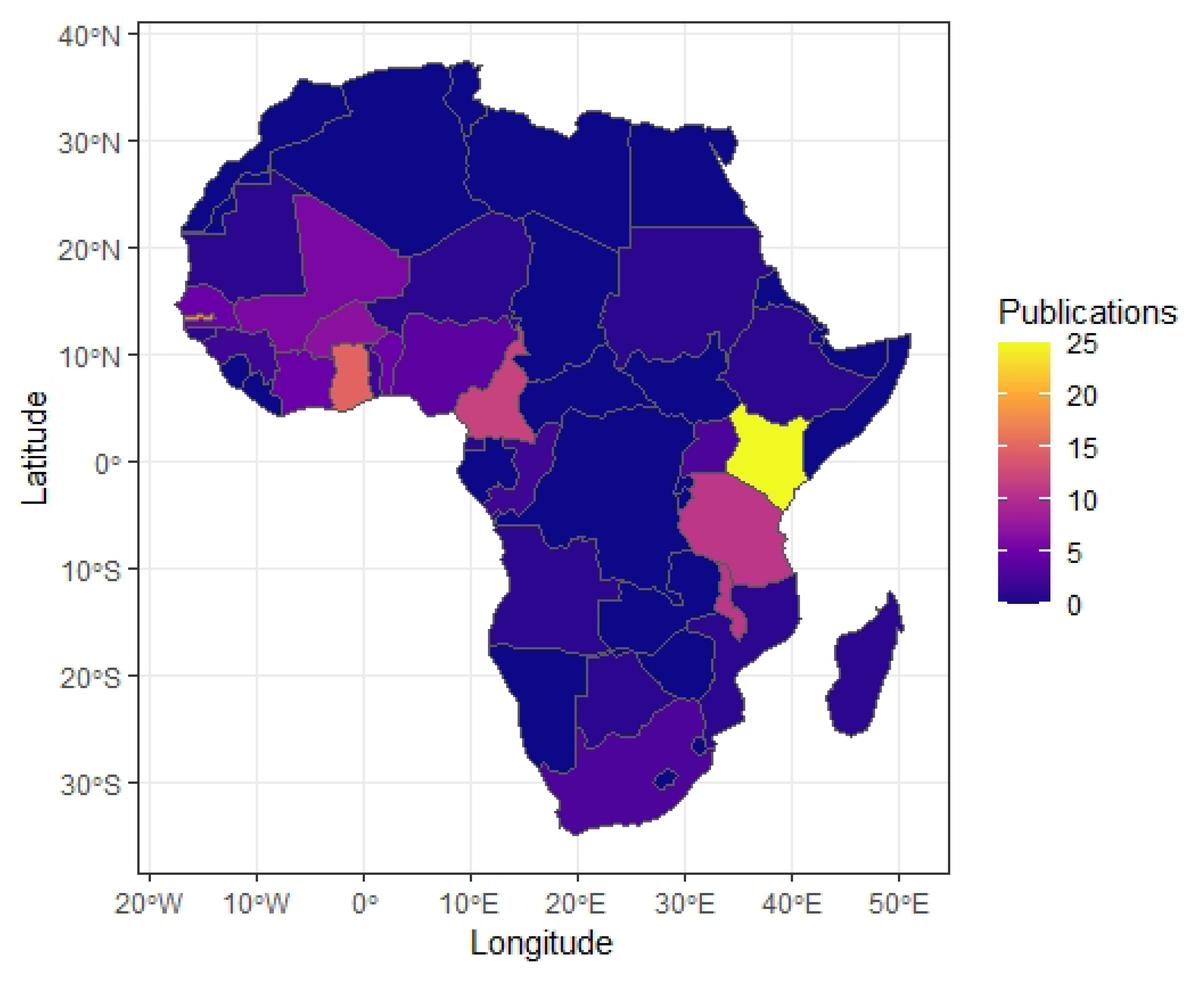
SSA countries conducting GWAS in malaria mapped with publications.

**Fig 6.**
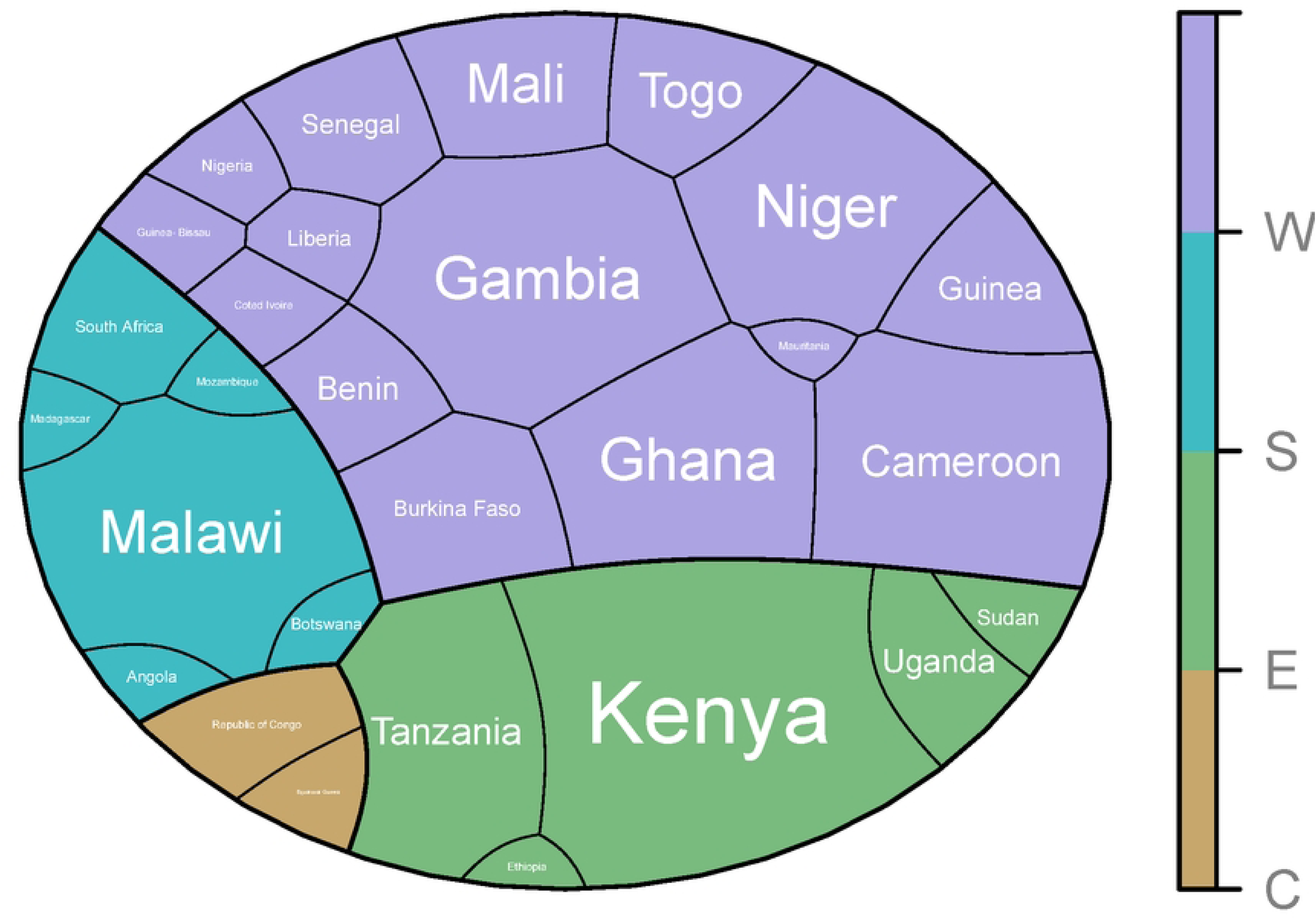
Polygon areas proportional to the number of studies conducted in SSA in different regions of Western Africa (W), Eastern Africa (E), Southern Africa (S), and Central Africa (C).

The research areas were further categorized into four distinct geographical regions namely, Western Africa (West), Southern Africa (Southern), Eastern Africa (Eastern), and Central Africa (Central) regions as shown in Fig 7. Many articles were mapped in the Western Africa region with Gambia having the highest number of articles (n=17).

**Fig 7.**
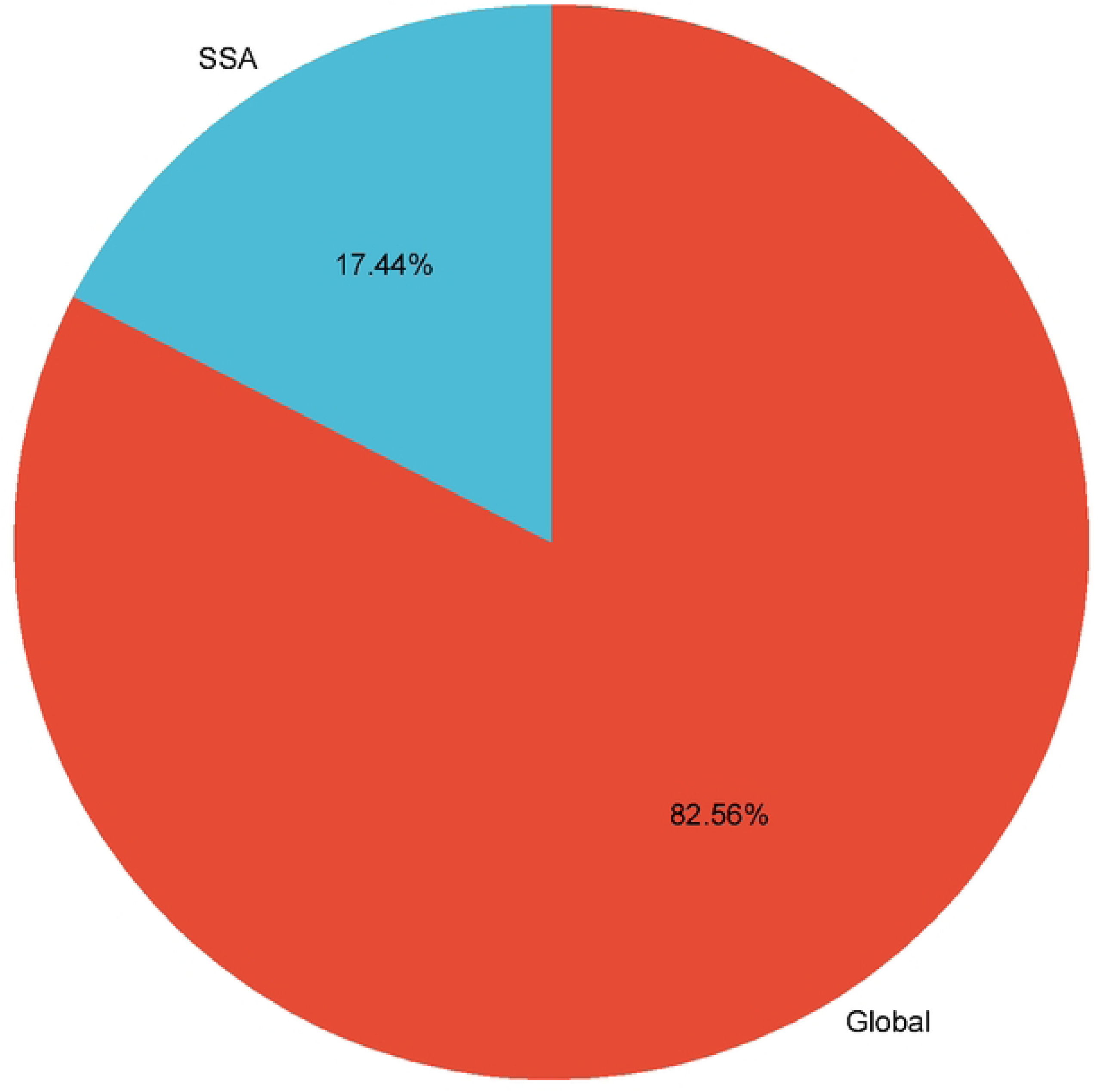
Percentage representation of GWAS research in malaria globally compared to SSA studies found on Google Scholar search between 2000 and 2023.

### Genetic variants and malaria

Most studies focused on genetic variants associated with susceptibility, severity and resistance to malaria (n=54) [2, 8–10, 26, 27, 29–38, 41, 42, 44–50, 52, 54–58, 61, 63–65, 67–69, 71, 74, 78–83]. Table 2 shows some single nucleotide polymorphisms (SNPs) and genes associated with malaria protection, and severity. The five studies [27, 37, 59, 63, 67] had genetic modes of inheritance on variants associated with malaria severity and protection. The studies are model-based, with heterosis, as one of the genetic inheritance modes associated with severe or mild malaria phenotypes. Ranvehall et al. [27] reported the most significant SNP association as the heterozygous protective rs334 significantly associated with resistance to severe disease. Manjurano et al. [37] reported the G6PD gene with heterozygous advantage effect which has a deficiency known to protect against severe malaria in the Tanzanian populations. Maiga et al. [63] reported some of the 19 regions of the genome under heterozygous advantage with significant SNPs associated with severe malaria. Other genetic models reported were the additive, recessive, and dominant modes in the studies.

**Table 2.**
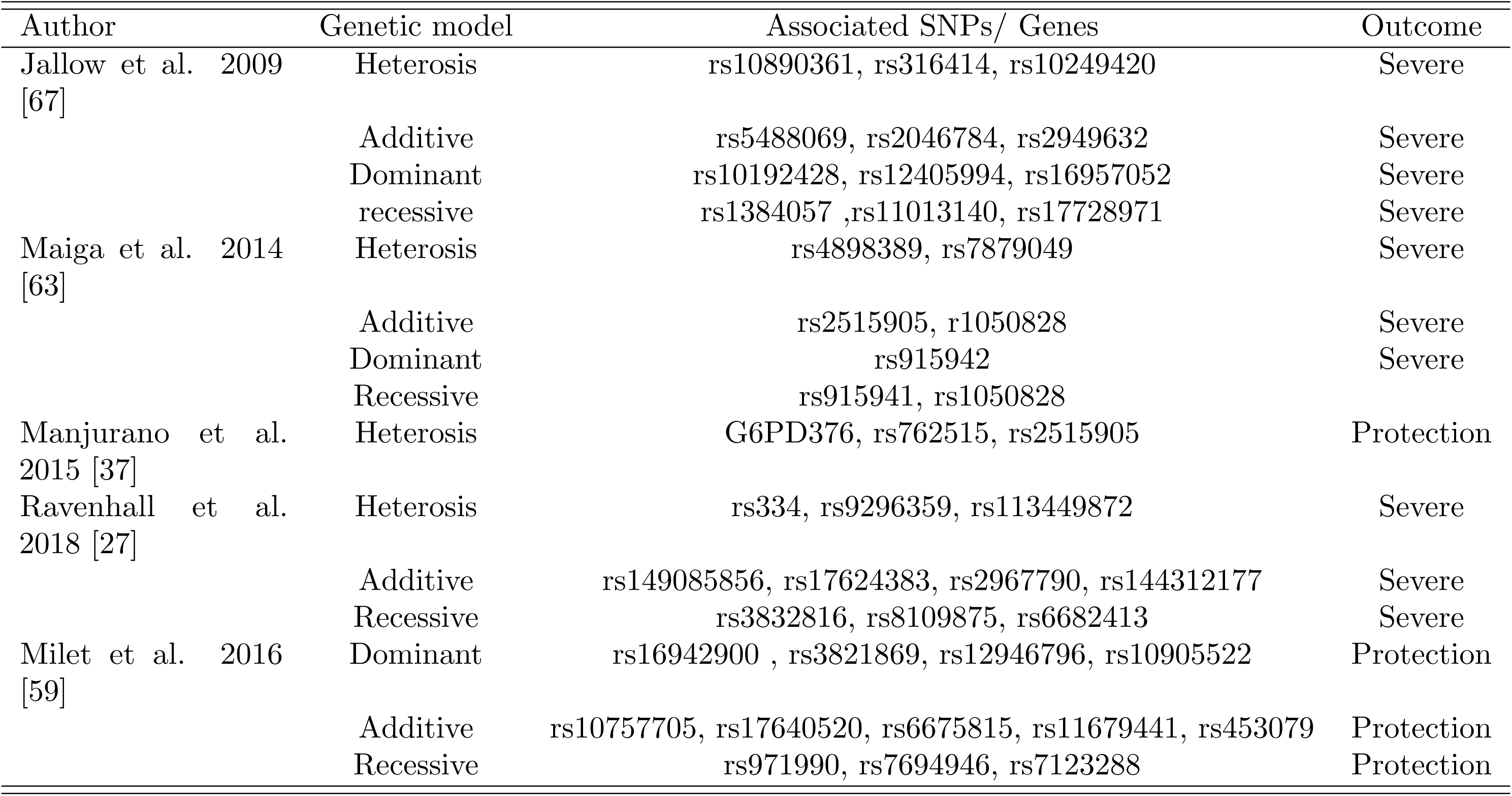
Associated SNPs and genes with protection, and severe malaria with underlying genetic models in the studies.

Studies on systematic reviews and meta-analysis ([8–10, 30, 64]), also included genetic models with studies by Kivinen et al. [30] including heterozygous mode of inheritance in the multi-center GWAS. Patterns of association with severe malaria were reported.

## Discussion

This scoping review aimed to identify existing malaria GWAS in SSA. A search was performed on Google Scholar, Pubmed, Scopus, and Web of Science and the results are highlighted in Fig 1. It included 99 articles with some studies covering topics in susceptibility, resistance, and severity of malaria, where genetic variants associated with malaria have been discovered and provided vital information. The number of studies conducted in SSA was significantly lower than the extensive global search for malaria GWAS, which produced 8,970 articles on Google Scholar (Fig 7). The research showed an increasing number of articles over time (Fig 2). In over 23 years, publications in malaria GWAS have consistently increased. The upward trend indicates that GWAS is increasingly recognized as a valuable tool for understanding the genetic factors that impact malaria susceptibility, resistance, and severity in SSA.

Most studies were conducted in Kenya and Gambia (Fig 5). The other countries with articles in malaria GWAS included Ghana, Cameroon, Tanzania, Burkina Faso, Benin, Uganda, Malawi, South Africa, Ethiopia, Ivory Coast, Senegal, Guinea, Congo, Angola, Mali, Sudan, Mauritania, Nigeria, Togo, Guinea, Mozambique, Equatorial Guinea and Madagascar. Despite being malaria-prone regions, some countries had no studies reported on the subject area. These included Zambia, Rwanda, Somalia, Chad, Namibia, Sierra Leone, Niger, Eritrea, Liberia, Burundi, Central African Republic, and Lesotho. Fig 6 further highlights the number of studies per region classified as Western, Eastern, Southern, and Central Africa. Many studies were reported in the Western region with Gambia having the highest number of publications in the region. Previous studies have uncovered more than 3 million genetic variants from African populations, some considered novel [17]. The diverse genetic variants have been observed among different ethnic communities; hence the potential to use knowledge to understand disease mechanisms and drug targets in clinical practices [16, 116]. Gouveia and colleagues [104] reported a high genetic diversity in Eastern Africa, while Ndo et al. [107] reported similar results in Western Africa. Although Africa is the continent with the highest genetic diversity, it has hosted only 2% of worldwide GWAS [17]. Studies on genetic diversity have shown that most GWAS focus mainly on European populations as highlighted in studies such as Sirugo et al. (2019) [117] and Melzer et al. (2020) [118]. Despite Africa’s significant genetic diversity, GWAS have not adequately been conducted in SSA.

Most study designs were case-control, accounting for 60% of the articles (Fig 3).

Case-control studies have been used for investigating disease risk factors and outcomes. They are less costly and less time-consuming in comparison to other study designs. In contrast to linkage studies, which require large sample sizes, case-control studies can detect genes that contribute only a minor fraction to the overall likelihood of a disease [119]. Heightened sensitivity is crucial in unraveling complex genetic associations. Studies on systematic reviews, meta-analyses, and other reviews were also noted. In addition, cohort, cross-sectional, and longitudinal designs were reported.

Many studies focused on susceptibility, severity, resistance, and associated genetic variants of malaria (Fig 4). Some studies reported SNPs and genes associated with protection and severe malaria outcomes under different genetic models (Table 2). Jallow et al. [67] reported regions with the strongest signal and associated models with the most significant SNPs found in dominant, trend, recessive, and heterozygous advantage models. Maiga et al. [63] reported significant SNPs found in dominant and heterozygous models associated with malaria phenotypes. Manjurano et al. [37] reported some associated SNPs found among female heterozygotes that offer protection against severe malaria. Other SNPs showed associations in additive, and recessive modes, as noted by Ravenhall et al. [27] in the Tanzanian population. The four Studies ([27, 37, 63, 67]) reported genetic variants, under heterotic mode of inheritance, associated with both severity and protection from malaria while studies by Milet et al. [59] conducted GWAS in children from Senegal and found more SNPs associated with protection against malaria under the additive, recessive, and dominant models. A recommendation to replicate such studies in other parts of Africa was suggested.

Furthermore, methodological tools, drug resistance patterns investigations, and intervention effectiveness evaluations emphasize the comprehensive nature of GWAS. For example, studies by Milet et al. [59] looked at the GWAS of antibody response to malaria vaccines while Ali et al. [53], studied the prevalence of drug resistance mutations among children in Cameroon. Several studies have used various statistical techniques, including Bayesian modeling [90] and generalized linear models [95], to explore genetic associations with malaria. Similar GWAS in other regions or those that address different phenotypes in SSA have adopted diverse methodologies [120–127], to investigate genetic associations with the respective phenotype. In addition, a range of GWAS software tools, such as *input2* as a framework used to impute genotypes, *bear* [93] and *R* [125], have been used to assess genetic correlations and explore associated genetic loci. A few studies on host-parasite interactions and evolutionary insights were also reported.

## Conclusion

The findings in genetic studies on malaria in SSA highlight the significant efforts put in to deepen our understanding of malaria. These efforts include an increasing number of publications, the use of various methodological approaches, research on a diverse population, and the study of malaria in different geographic locations. However, despite the considerable genetic diversity observed in SSA, global representation in research volume still needs to be achieved. There is also a lot of untapped potential to develop novel methodologies and discover more genetic loci related to susceptibility, resistance, and severity of malaria.

The African Genome Variation project and the Africa BioGenome projects, among other projects, have enabled genetic disease studies in SSA by discovering millions of genetic variants [128, 129]. Such projects provide platforms for collaborations and partnerships among African scientists through biodiversity genomics.

## Supporting information

*R* software and *SRplot* platform for data visualization and graphing were used.

## Data Availability

All relevant data are within the manuscript and its Supporting Information files.

## Acknowledgments

We acknowledge and express our gratitude to Strathmore University for their support during this research.

